# Social distance and SARS memory: impact on the public awareness of 2019 novel coronavirus (COVID-19) outbreak

**DOI:** 10.1101/2020.03.11.20033688

**Authors:** Haohui Chen, Weipan Xu, Cecile Paris, Andrew Reeson, Xun Li

## Abstract

This study examines publicly available online search data in China to investigate the spread of public awareness of the 2019 novel coronavirus (COVID-19) outbreak. We found that cities that suffered from SARS and have greater migration ties to the epicentre, Wuhan, had earlier, stronger and more durable public awareness of the outbreak. Our data indicate that forty-eight such cities developed awareness up to 19 days earlier than 255 comparable cities, giving them an opportunity to better prepare. This study suggests that it is important to consider memory of prior catastrophic events as they will influence the public response to emerging threats.

## Introduction

Public awareness is important in managing the spread of infectious diseases. Individual actions, such as increased attention to hygiene and avoiding crowds, can reduce disease spread. Awareness also supports rapid identification and treatment of new cases and facilitates collective responses, such as closures of schools or transit systems^1^. In the modern world, diseases can move faster than ever due to the growing movement of people between cities, regions and countries. However, digital technology means information can move even faster, providing an opportunity for individuals and communities to protect themselves ahead of the disease itself arriving.^2^ This study considers the spread, and persistence, of public awareness of the novel Wuhan coronavirus which emerged in late 2019. During the first few weeks of this outbreak there was little coverage from mainstream media outlets, providing an unusual opportunity to study the spread of awareness of an emerging disease via other channels.

Previous studies show that the spread of awareness is strongly related to the physical locations of individuals in a social network in relation to the unfolding events^2–4^, termed the *social distance* effect. In online social networks, people with more connections tend to receive earlier warnings of catastrophic events. For example, in Hurricane Sandy in the USA, Twitter users with more followers had an awareness lead-time of up to 26 hours than less connected users^4^. Moreover, the magnitude of awareness increases over decreasing distances to the epidemic centers. For example, public awareness in Weibo, a Chinese social media platform, was two orders of magnitude stronger for the H7N9 influenza outbreak that occurred in China than the Middle East Respiratory Syndrome Coronavirus (MERS-CoV) outbreak that occurred elsewhere^5^.

Experience of similar events, such as outbreaks of H5NI influenza in 2001, SARS (Severe Acute Respiratory Syndrome) in 2003, H1N1 influenza in 2009 and Ebola in 2014 is also likely to influence awareness. In China, the outbreak of SARS between 2003 and 2004 caused a total of 7,429 reported cases and 685 deaths^6^, and had a lasting traumatic impact on survivors and communities ^7,8^. In this work, we set out to test whether public awareness of the new disease outbreak is related to social distance from the centre of the epidemic and past experience of the SARS epidemic in 2003. The SARS outbreak was 17 years ago, but its horror might still condition public awareness of lethal infectious diseases. To the best of our knowledge, few studies were carried out to understand how past severe outbreaks affect public awareness when a new outbreak occurs. This study estimates the post-SARS effect, called *SARS memory* effect, on the current outbreak.

We use the continuing Wuhan coronavirus outbreak as our case study to estimate the effects of *social distance* and *SARS memory* on the spread of public awareness. In late 2019, a new coronavirus, designated as COVID-19, was identified in Wuhan, the capital of China’s Hubei province^9^. As of February 1st, 2020, the virus has caused approximately 11,184 cases and 258 deaths in 21 countries with the majority of cases in mainland China^10^. The first symptoms were reported on December 1st, 2019^11^, but there was no solid evidence of human-to-human transmission until January 10^th^, 2020, when a patient, who did not travel to Wuhan, became infected with the virus after several days of contact with four family members^12^. However, there was little information available to the public until an official announcement about human-to-human transmission of the virus on January 20^th^, 2020^13^. Wuhan City, thoroughfare (“*九省通衢*”), is in the central part of China, serving as a major transportation hub transiting more than 120 million passengers every year^14^. The massive numbers of transits provided a perfect opportunity for the virus to spread. Another feature is the timing of the outbreak, close to the Spring Festival travel season, *Chunyun* (“*春运*”), which started on January 10^th^, 2020. While the virus spread across almost all provinces, 16 cities had been locked down by January 23^rd^, 2020^15^. Accordingly, this study focuses on the time period between December 15^th^, 2019 and Jan 23^rd^, 2020.

## Data and methods

### Public awareness measurement

Seeking epidemic-related information online can provide an indicator of public awareness of this new disease. In this study, we use the Baidu Search Index (BSI), available publicly at http://index.baidu.com, to measure the public awareness over time and locations (e.g., city). The total number of internet users using the Baidu search engine reached 649 million in 2014, accounting for 47.9% of the national population^16^. BSI has been used to predict epidemic outbreak^17^, HIV/AIDS incidence^18^ and tourism flows^19^, suggesting BSI can provide a representative proxy for public awareness. BSI provides a weighted index for each search term. In this study, we used the term “Wuhan pneumonia” (“*武汉肺炎*”), as the public has widely used. We also tried “novel coronavirus” (“*新型冠状病毒*”) and “Wuhan outbreak” (“*武汉爆发*”), but the former did not exhibit a search surge, and the latter was not indexed by Baidu. Due to the privacy concern, Baidu masks daily readings that are below 57 as zero. Therefore, we used the maximum BSI value of the search term “common cold” (“*感冒*”) between Dec 10^th^ and 31^th^, 2019, to control the size effect for each city. We use the Ljung-Box test^20^ to estimate whether or not the daily readings of “common cold” are stationary. As a result, the daily readings of 18 out of 364 cities were found to be non-stationary, so they were excluded from this study. The magnitude of public awareness of the Wuhan outbreak over time t and city i ∈ {1, …, 346} can be represented as 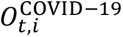 as defined below. 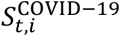 and 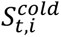 represent the BSI values of the search terms “Wuhan pneumonia” and “common cold” respectively. The BSI raw data is provided in S1 of the supplementary materials (SM).

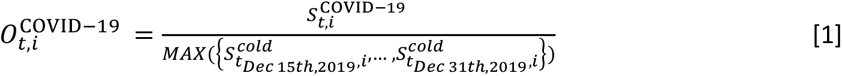

The earliest day the magnitude of public awareness exceeds the arbitrary thresholds *C* ∈ {1.5, 2, 3, 4} is defined as the earliest warning day, *t*_*warming*(*i*)_, for city i. We also define the starting day of *Chunyun* as *t*_*chunyun*_, indicating the onset day when it is likely the virus would reach all cities. As *Chunyun* transited approximately 3 billion million passengers in 40 days in 2019^21^, crowded transport hubs create perfect opportunities for the virus to spread. Therefore, the earlier the lead-time awareness, the better for infection control. The lead-time of awareness for city i is thus defined as:

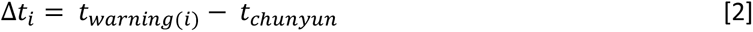

Awareness typically follows a cyclical process, called the unaware-aware-unaware (UAU) process, as time passes. Keeping the public at a high level of awareness could help mitigate the virus transmission process. Therefore, we also measure the awareness retention rate as the average of magnitude from the next day of *t*_*warming*(*i*)_ to the day of *Chunyun* over the magnitude at *t*_*warming*(*i*)_.

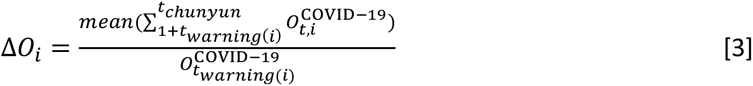

### Measuring social distances

Microblogs (e.g., Weibo) and private social media (e.g., WeChat) are the primary communication tools used by most Chinese people^16^. While information flows cannot be observed directly, empirical studies show that social networks are influenced by long distance travel.^22^ We therefore use migration flows as a proxy for long-distance information flows To be more specific, if workers born and raised in city A now work in city B, they are likely to relate information about an epidemic in city B back to friends and family in city A. This is particularly relevant in the Chinese context, where migrant workers account for more than one-third of the working population^23^.

We use the migration flows extracted from the Baidu Migration Matrix (BMM) to build a migration network (Fig. 1). We then compute the shortest steps between any city to Wuhan, deriving the variable *social distances* for city i as *D*_*i*_ ∈ (1, 8). Wuhan and the cities located in Hubei province have *D*_*i*_ = 1, while cities located far away from Wuhan tend to have larger values, e.g., *D*_*i*=*Lhasa*_ = 6.

**Figure 1.**
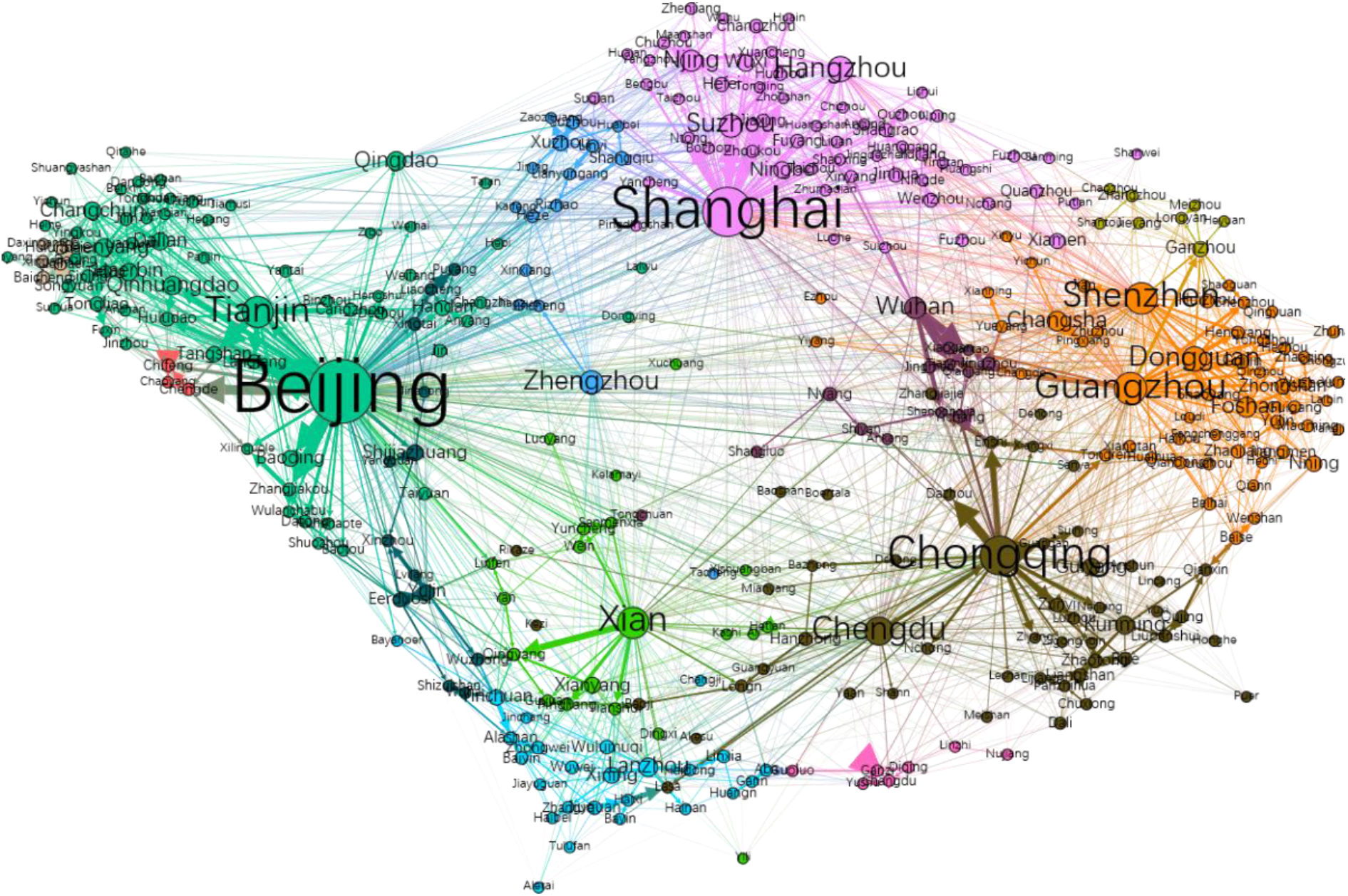
The migration network based on Baidu Migration Matrix. The nodes represent cities in mainland China, and the length and thickness of the edges indicate the travel frequency between the corresponding pair of cities. The node size is proportional to the numbers of transit passengers at the corresponding city. The network does not include Hong Kong, Macau and Taiwan, as the Baidu Migration Matrix does not include them.

Moreover, we added *D*_*i*=*Hong Kong*_ = 1, based on the rationale that Hong Kong has more airline traffic flows to Wuhan than Shanghai^24^, which has *D*_*i*=*Shanghai*_ = 1. Macau and Taiwan both have frequent traffic flows to Hong Kong^24^, so we added *D*_*i*=*Macau*_ = 2 and *D*_*i*=*Taiwan*_ = 2.

### Measuring SARS memory

We collected all reported SARS cases in mainland China, Hong Kong, Taiwan and Macau, and assign the numbers of cases to each city as *SARS*_*i*_, *i* ∈ {1, …, 346}. The values range between zero (no reported cases) and 2,521 (*i* = *Beijing*), with an average of 70.8 and a median of 4 cases in cities with at least one case reported. We then use the logarithm, as most cities reported zero cases of SARS resulting in:

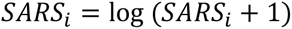

### Estimating social distance and SARS memory effects

We build three groups of regression models to estimate the effects of *social distance* and *SARS memory* on public awareness measures 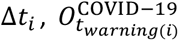 and Δ*O*_*i*_ respectively. *GDP_per_capita*_*i*_represents the gross domestic product (GDP) per capita for city *i. SubProvincial*_*i*_ indicates whether or not city *i* has sub-provincial or greater administrative power. Sub-provincial cities are mostly capitals of the provinces in which they are located, or important cities designated by the central government. Four cities, including Beijing, Shanghai, Tianjin and Chongqing, which are under direct control of the central government are also labelled as *SubProvincial*_*i*_ = 1. Those sub-provincial and above cities have much better facilities and expertise for infection control than other cities^25^, so we assume residents could be more alert. *SubProvincial*_*i*_ is used to control the effects of *administrative level*. We also introduce Euclidean distances as a control variable, denoted as *d*_*i*_.

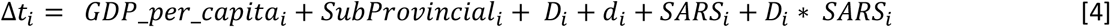

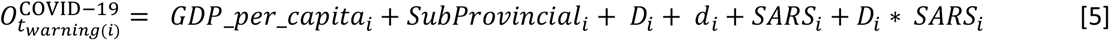

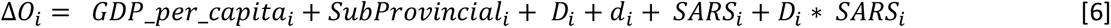

All data necessary to replicate the analysis is attached as S2 of the SM.

## Results

### The early warnings of the outbreak

As early as Dec 31^st^, 2019, most of the cities (326 out of 346) have exhibited at least some awareness of the emerging Wuhan outbreak (Fig. 2B). However, awareness then decreased until Jan 19^th^, 2020, one day before the Chinese Centre for Disease Control and Prevention confirmed human-to-human transmissions of the novel coronavirus. Since Jan 20^th^, 2020, overall awareness has increased by a magnitude of at least five, demonstrating significant awareness across all cities (Fig. 2B). Awareness remained low as the epidemic spread, falling close to its lowest point on the starting day of *Chunyun* (Jan 10^th^, 2020). Considering cities that showed initial novel coronavirus awareness levels at least 1.5 times that of the search term “common cold”, we found a total of 166 alert cities as early as Dec 31^st^, 2019 (48 cities at a tighter threshold of *C* = 3.0 times, illustrated in Fig. 2A). However, awareness decreased significantly during *Chunyun*.

**Figure 2.**
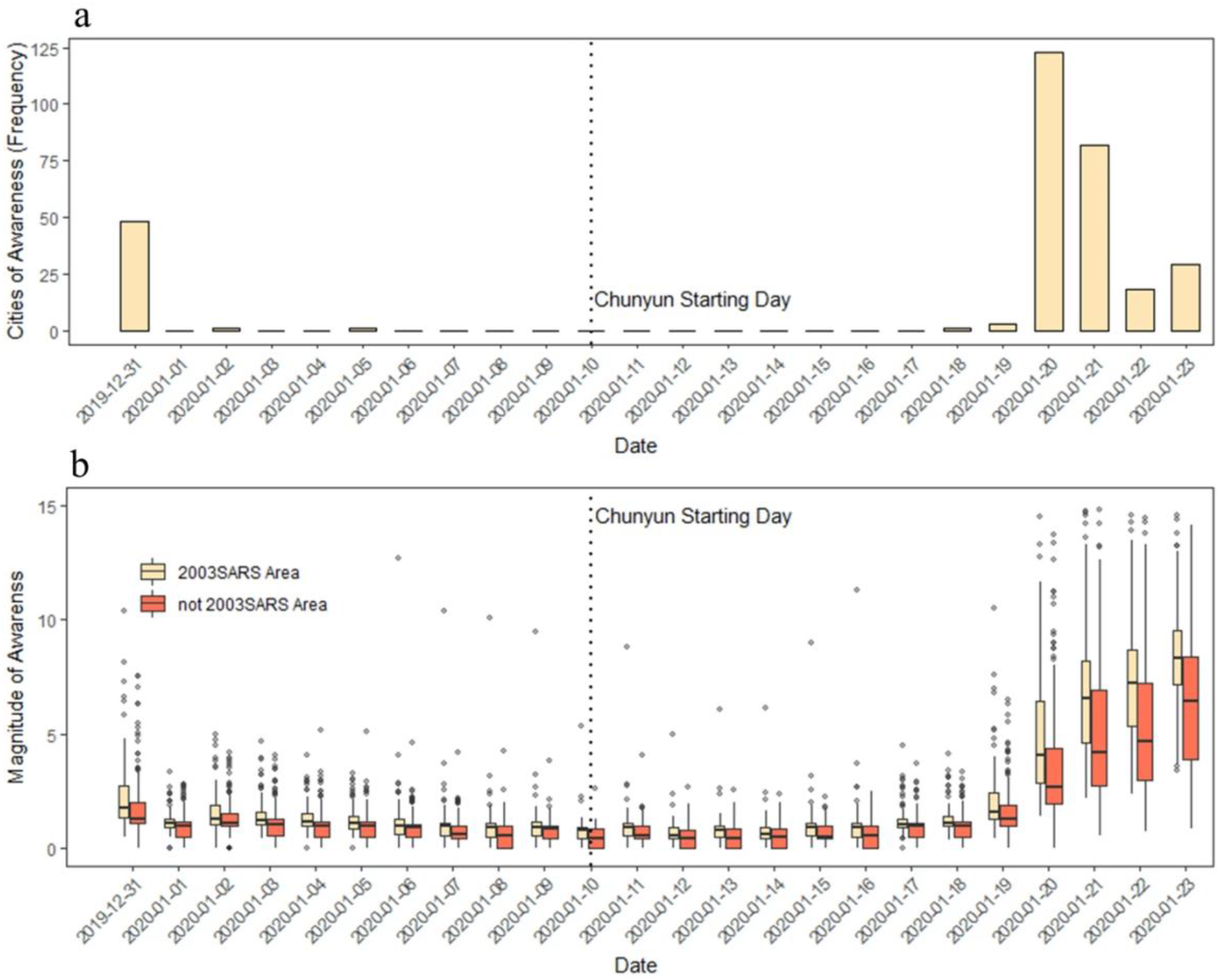
Public awareness over time. a) the frequency distributions of cities that exhibit the first significant signal of awareness over time. The number of cities for which searches for “Wuhan pneumonia” exceed *C* = *3* times the search term “common cold” is reported every day. b) public awareness on the topic of “Wuhan pneumonia” over time. All 346 cities exhibit at least some searches of the term “Wuhan pneumonia” during the initial outbreak period. Of these, 326 cities recorded searches about it as early as Dec 31st, 2019. Cities are divided into two groups according to whether or not they had reported SARS cases in 2003-04.

The evolution of public awareness over time followed an unusual pattern. In a typical UAU process, people are unaware of emerging catastrophic events until they are told by their social contacts. They remain aware during the event, and awareness then fades subsequently^2,26^. However, during the Wuhan outbreak, the public experienced a process as aware-unaware-aware, with public awareness declining during the early phase of the outbreak.

Dividing cities into two groups of equal size according to the numbers of reported SARS cases, we found the cities that had been struck by SARS to be more alert (p < 0.05) during onset (2B). Therefore, we believe the *SARS memory* still conditions public awareness. We provide evidence of its effects at the end of this section.

### Awareness advantage

Three features define the awareness advantage of *alert* cities, including early awareness, strong magnitude and high retention of awareness.

#### Lead-time advantage

During onset, between Dec 31^st^, 2019 and Jan 23^rd^, 2020, 266 cities exhibited significant public awareness (using a threshold at *C* = 3.0; 210 cities at *C* = 4.0; 314 cities at *C* = 2.0; and 322 cities at *C* = 1.5). The lead-time advantage Δ*t*_*i*_ ranges between −13 and 10 (*C* = 3.0), with an average of −7.4 days and a median of −10 days. Forty-eight cities emerge with early signals of public awareness, as early as Dec 31^st^, 2019, while for most others (255 cities), awareness is as late as Jan 20^th^, 2020 (Fig. 2A). The cities of substantial lead-time advantage are those either closed to Wuhan in terms of social distances *D*_*i*_ or struck by the SARS outbreak. For example, Changchun in Jilin province with 34 SARS cases and far away from Wuhan still achieved a ten days lead-time advantage. The cities that did not exhibit awareness, such as Qaramay and Heihe, are mainly located far away from Wuhan and did not suffer from the SARS outbreak.

#### Magnitude of awareness

In terms of the magnitude of awareness, all 346 cities exhibit at least some awareness during the onset. The values of 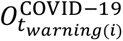 range between 0.50 and 61.51, with an average of 2.18 and a median of 1.49. Wuhan undoubtedly ranked the first, while Wuzhong in Ningxia ranked last. Cities in Hubei province exhibit much greater awareness, with an average of 7.83 and a median of 3.46. Shenzhen, Shanghai and Beijing also had high scores at 10.40, 10.65, and 9.78 respectively. Those three cities are both close to Wuhan in terms of social distances *D*_*i*_ and struck by SARS.

#### Retention of awareness

Even though most of the cities exhibit at least some awareness as early as Dec 31^st^, 2019, only a few retain it over the following weeks as the virus began to spread. The retention rates, Δ*O*_*i*_, range between zero and 137%, with an average of 54% and a median of 55%. Eight cities lost awareness before *Chunyun*, while four cities developed greater awareness. Xilingol League in Inner Mongolia ranked 4^th^, with a retention rate at 103%. Xilingol is far away from Wuhan in terms of social distance, but it was struck by SARS. It is worth noting that a confirmed case of plague was reported in Xilingol on Nov 16^th^, 2019, only 45 days before the Wuhan outbreak.

#### Estimation of the social distance and SARS memory effects

The effects of *social distance* and *SARS memory* on the lead-time advantage are estimated according to Eq. 4, controlled by Euclidean distances, GDP per capita and the city’s *administrative level* (Table 1). We found that, in model (3) in Table 1, *SARS*_*i*_ exhibits positive effects, while *D*_*i*_ shows a negative association with awareness. That means cities of strong *SARS memory* and which are closer to Wuhan in terms of *Social distances* develop early awareness. Moreover, the interaction term *D*_*i*_ ∗ *SARS*_*i*_ exhibits negative effects, indicating that the SARS *memory* effect becomes stronger where cities are closer to Wuhan in terms of *social distances*.

**Table 1.** Estimate social distance and SARS memory effects on lead-time advantage.

| Dependent variable: |  |
| --- | --- |
| Lead-time $\Delta t_i$ ($C = 3.0$) | |

|  | (1) | (2) | (3) | (4) | (5) | (6) | (7) |
| --- | --- | --- | --- | --- | --- | --- | --- |
| $SARS_i$ | 1.916***<br>(0.345) | | 2.566***<br>(0.664) | | 4.644<br>(4.531) | 1.590***<br>(0.593) | -2.276<br>(4.016) |
| Social distance $D_i$ | | -3.222***<br>(0.271) | -2.753***<br>(0.286) | | -2.310***<br>(0.372) | -1.476***<br>(0.315) | -1.367***<br>(0.334) |
| $D_i * SARS_i$ | | | -0.617***<br>(0.238) | | -0.660***<br>(0.243) | -0.412**<br>(0.206) | -0.453**<br>(0.211) |
| $\log(d_i)$ | | | | -3.567***<br>(0.527) | -1.125<br>(0.720) | -1.744***<br>(0.490) | -2.139***<br>(0.637) |
| $\log(d_i) * SARS_i$ | | | | | -0.267<br>(0.679) | | 0.583<br>(0.599) |
| $SubProvincial_i$ | | | | | | 8.516***<br>(1.135) | 8.763***<br>(1.163) |
| $GDP\_per\_capita_i$ | | | | | | 2.818***<br>(0.599) | 2.762***<br>(0.602) |
| Constant | -8.571***<br>(0.470) | 3.766***<br>(1.007) | 1.610<br>(1.104) | 16.287***<br>(3.523) | 7.441*<br>(4.121) | -22.169***<br>(7.068) | -19.291**<br>(7.662) |
| Observations | 306 | 306 | 306 | 306 | 306 | 305 | 305 |
| R<sup>2</sup> | 0.092 | 0.318 | 0.357 | 0.131 | 0.368 | 0.535 | 0.537 |
| Adjusted R<sup>2</sup> | 0.089 | 0.316 | 0.351 | 0.128 | 0.358 | 0.526 | 0.526 |
| Residual Std. Error | 7.356 (df = 304) | 6.376 (df = 304) | 6.211 (df = 302) | 7.196 (df = 304) | 6.176 (df = 300) | 5.312 (df = 298) | 5.312 (df = 297) |
| F Statistic | 30.820*** (df = 1; 304) | 141.645*** (df = 1; 304) | 55.885*** (df = 3; 302) | 45.859*** (df = 1; 304) | 34.999*** (df = 5; 300) | 57.218*** (df = 6; 298) | 49.170*** (df = 7; 297) |

Note:
\*p\*\*p\*\*\*p<0.01

While controlling the model with Euclidean distances (model (5) in Table 1), we found that *SARS memory* effect becomes non-significant, but *social distance* and its interaction with *SARS memory* hold. Meanwhile, Euclidean distances effect are non-significant, even though it exhibits negative effect alone in model (4) in Table 1.

We further control the model with GDP per capita and administrative level (model (6) & (7) in Table 1). Model (6) is more favorable than model (7), because the former achieves slightly lower AIC scores (Akaike’s Information Criteria)^27^ at 1893.132 with fewer degrees of freedom (df = 8) than the latter (AIC = 1894.160, df = 9). In model (6) in Table 1, we found that both *social distance* and Euclidean distances exhibit negative effects, but the *social distance* effects decrease almost half compared to model (5) in Table 1. The *SARS memory* effects hold. Also, the interaction term *D*_*i*_ ∗ *SARS*_*i*_ is still significant, which means cities of stronger *SARS memory* will develop more lead-time advantage, particularly when they are closer to Wuhan. GDP per capita and the binary variable *SubProvincial*_*i*_ exhibit significant positive effects on the lead-time advantage.

The effects of *social distance* and *SARS memory* on the magnitude of awareness are estimated according to Eq. 5 (Table 2). Similar to the findings in Table 1, *SARS*_*i*_ memory positively affects public awareness in all models. *Social distances D*_*i*_ show a significant negative effect only in the models without controlling variables (model (1), (2) & (3) in Table 2). However, the interaction term between *social distances* with *SARS memory* show a significant negative effect. When we control by Euclidean distances, GDP per capita and the administrative level (model (5) and (6) in Table 2), those effects still hold. Moreover, the effects of administrative level and development level both exhibit positive effects on the magnitude of awareness. We hypothesize that residents with better education (proxied by GDP per capita) better understand the danger of deadly infectious diseases and, accordingly, tend to seek up-to-date information online.

**Table 2.** Estimate social distance and SARS memory effects on the Magnitude of awareness at the earliest day of awareness.

|  | Dependent variable: |  |  |  |  |  |
| --- | --- | --- | --- | --- | --- | --- |
| | Magnitude of awareness at the earliest day of awareness: $O_{t_{warning(t)}}^{19-nCoV}$ | | | | | |
|  | (1) | (2) | (3) | (4) | (5) | (6) |
| $SARS_i$ | 0.550***<br>(0.163) | | 1.308***<br>(0.350) | | 25.629***<br>(1.539) | 23.752***<br>(1.432) |
| Social distance $D_i$ | | -0.791***<br>(0.126) | -0.619***<br>(0.133) | | -0.100<br>(0.116) | 0.164<br>(0.110) |
| $D_i * SARS_i$ | | | -0.412***<br>(0.125) | | -0.284***<br>(0.082) | -0.218***<br>(0.075) |
| $\log(d_i)$ | | | | -2.534***<br>(0.204) | -0.730***<br>(0.240) | -0.988***<br>(0.222) |
| $\log(d_i) * SARS_i$ | | | | | -3.577*** | -3.354*** |
|  |  |  |  |  | (0.231) | (0.214) |
| *SubProvincial<sub>i</sub>* |  |  |  |  |  | 2.438*** |
|  |  |  |  |  |  | (0.415) |
| *GDP_per_capita<sub>i</sub>* |  |  |  |  |  | 0.961*** |
|  |  |  |  |  |  | (0.206) |
| Constant | 1.874***<br>(0.211) | 5.071***<br>(0.495) | 4.342***<br>(0.546) | 19.208***<br>(1.381) | 6.933***<br>(1.370) | -2.702<br>(2.643) |
| Observations | 338 | 338 | 338 | 338 | 338 | 336 |
| R<sup>2</sup> | 0.033 | 0.106 | 0.141 | 0.315 | 0.653 | 0.720 |
| Adjusted R<sup>2</sup> | 0.030 | 0.103 | 0.134 | 0.313 | 0.648 | 0.714 |
| Residual Std. Error | 3.518 (df = 336) | 3.382 (df = 336) | 3.324 (df = 334) | 2.961 (df = 336) | 2.120 (df = 332) | 1.915 (df = 328) |
| F Statistic | 11.384*** (df = 1; 336) | 39.693*** (df = 1; 336) | 18.349*** (df = 3; 334) | 154.271*** (df = 1; 336) | 124.843*** (df = 5; 332) | 120.568*** (df = 7; 328) |

Note:
\*p\*\*p\*\*\*p<0.01

The effects of *social distance* and *SARS memory* on retention of awareness are estimated according to Eq. 6. Unlike the results in Tables 1 and 2, we observe no effects from *SARS memory* (model (6) in Table 3). When we control Euclidean distances, the development level and the administrative level, the explanatory power of the model is still relatively weak (Adj. *R*^*2*^ = 0.104). It seems the decreasing awareness is a collective behavior that occurred simultaneously. Interestingly, *social distances* have a significant effect while the Euclidean distances do not. Development level exhibits positive effects, which suggests residents of better educated cities could be more alert during the epidemic onset. However, administrative level shows a negative effect. It seems residents living in important cities (in terms of administrative power) lost interest in the infectious diseases before *Chunyun*.

**Table 3.**
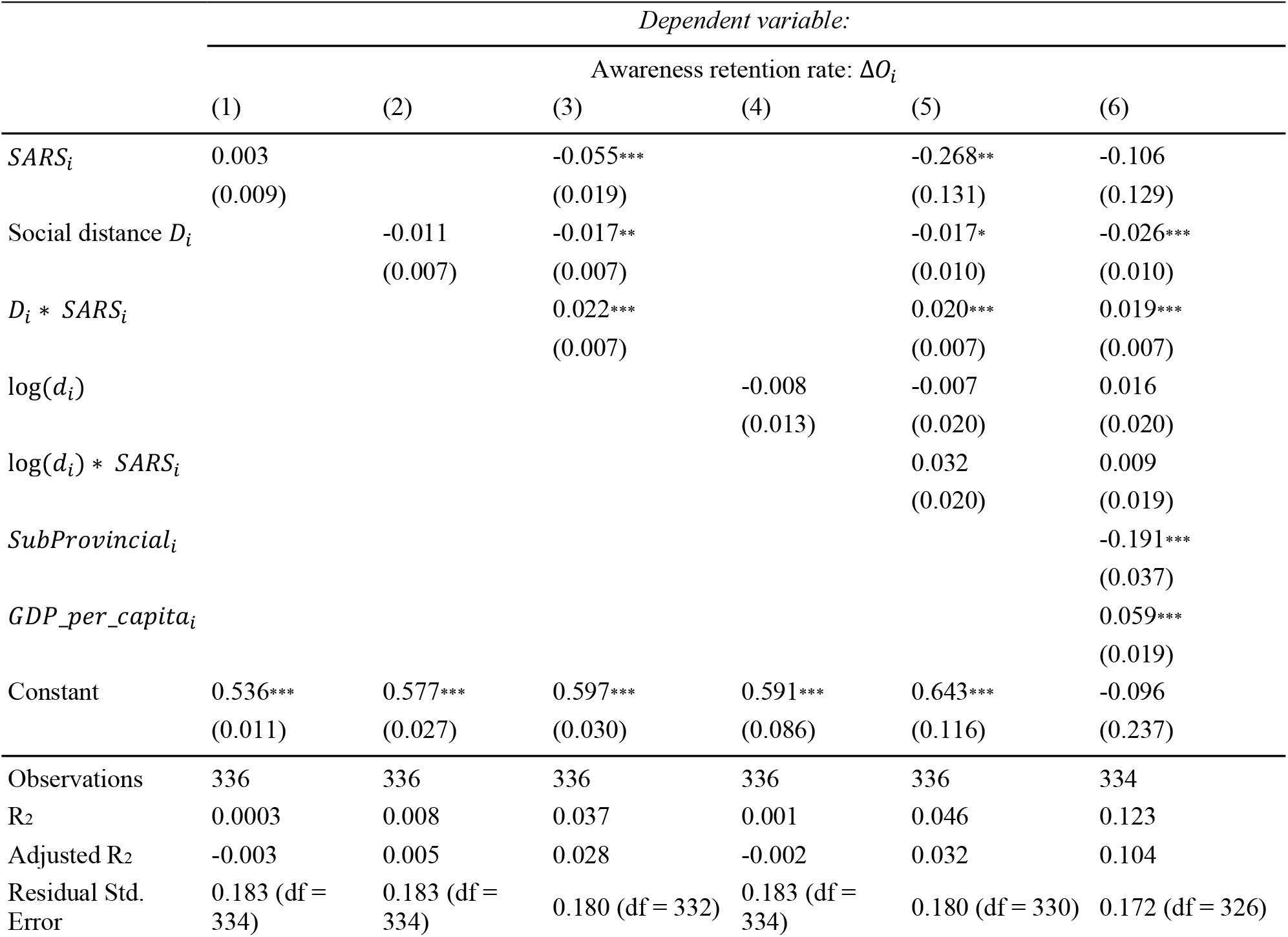
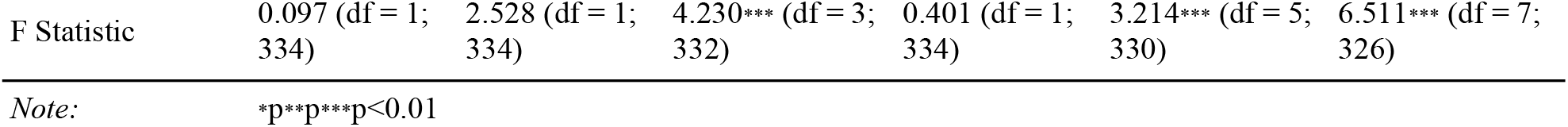
Estimate social distance and SARS memory effects on the awareness retention rate.

|  | Dependent variable: |  |  |  |  |  |
| --- | --- | --- | --- | --- | --- | --- |
| | Awareness retention rate: $\Delta O_i$ | | | | | |
|  | (1) | (2) | (3) | (4) | (5) | (6) |
| *SARS<sub>i</sub>* | 0.003<br>(0.009) |  | -0.055***<br>(0.019) |  | -0.268**<br>(0.131) | -0.106<br>(0.129) |
| Social distance *D<sub>i</sub>* |  | -0.011<br>(0.007) | -0.017**<br>(0.007) |  | -0.017*<br>(0.010) | -0.026***<br>(0.010) |
| *D<sub>i</sub> \* SARS<sub>i</sub>* |  |  | 0.022***<br>(0.007) |  | 0.020***<br>(0.007) | 0.019***<br>(0.007) |
| $\log(d_i)$ | | | | -0.008<br>(0.013) | -0.007<br>(0.020) | 0.016<br>(0.020) |
| $\log(d_i) * SARS_i$ | | | | | 0.032<br>(0.020) | 0.009<br>(0.019) |
| *SubProvincial<sub>i</sub>* |  |  |  |  |  | -0.191***<br>(0.037) |
| *GDP_per_capita<sub>i</sub>* |  |  |  |  |  | 0.059***<br>(0.019) |
| Constant | 0.536***<br>(0.011) | 0.577***<br>(0.027) | 0.597***<br>(0.030) | 0.591***<br>(0.086) | 0.643***<br>(0.116) | -0.096<br>(0.237) |
| Observations | 336 | 336 | 336 | 336 | 336 | 334 |
| R<sup>2</sup> | 0.0003 | 0.008 | 0.037 | 0.001 | 0.046 | 0.123 |
| Adjusted R<sup>2</sup> | -0.003 | 0.005 | 0.028 | -0.002 | 0.032 | 0.104 |
| Residual Std. Error | 0.183 (df = 334) | 0.183 (df = 334) | 0.180 (df = 332) | 0.183 (df = 334) | 0.180 (df = 330) | 0.172 (df = 326) |
| F Statistic | 0.097 (df = 1; 334) | 2.528 (df = 1; 334) | 4.230*** (df = 3; 332) | 0.401 (df = 1; 334) | 3.214*** (df = 5; 330) | 6.511*** (df = 7; 326) |

Note: \*p\*\*p\*\*\*p<0.01

## Discussion

The Wuhan coronavirus outbreak is still striking China, with thousands of cases and dozens of deaths reported every day. From this study, we found that the spread of public awareness varies markedly across Chinese cities. Through controlling for development, administrative levels, and Euclidean distances, we observe cities that were struck by SARS and have more migration to the epicentre, Wuhan, showed earlier, stronger and more durable public awareness of the outbreak. Moreover, 48 cities had developed public awareness as early as Dec 31^st^, 2019, with up to 19 days of lead-time advantage against some other 255 cities. The study suggests that memory of previous events, as well as social links to an emerging threat, may influence public behaviour. Greater awareness could help slow the spread of a disease, for example through increased attention to hygiene, mask-wearing and reduced interpersonal contact. It might also facilitate collective responses such as enforced quarantine measures. However, in some circumstances enhanced awareness could have negative impacts, such as unnecessary panic or ostracism of groups perceived as being at greater risk of infection

Due to the lack of infection statistics, we cannot yet statistically estimate the effect of public awareness on the subsequent seriousness of the outbreak. We note that Xilingol League in Inner Mongolia, which had relatively stronger and more durable public awareness, had fewer cases (two cases as reported at Feb 7^th^, 2020^10^) than other cities in the same province (totally 50 cases, with an average of 4.55 cases per city^10^).

To the best of our knowledge, this study is the first to investigate how memory of previous catastrophic events, e.g., SARS, and *social distances* could affect the spread of public awareness. Further studies will be needed to understand whether this holds in other context, beyond the unusual and tragic circumstances of the noel coronavirus in Wuhan.

## Data Availability

All data needed to evaluate the conclusions in the paper are present in the paper and/or the Supplementary Materials.

## Acknowledgments

We thank Dr. Ross Sparks from Data61, CSIRO for helpful suggestions and advice on the methodologies. We also thank the anonymous reviewers for their valuable suggestions.

## Author contributions

X.L., H.C., C.P. and A.R. conceived of the research question. H.C. and W.X. processed the Baidu Search Index data. X.L. and W.X. collected the other meta data including SARS cases and GDP per capita. H.C., X.L., W.X., A.R. and C.P. interpreted the result. H.C., X.L., W.X., A.R. and C.P. drafted the manuscript and compiled supplementary information. All authors edited the manuscript and supplementary information and aided in concept development.

## Supplementary Materials

File S1: Baidu Search Index data

File S2: The data necessary to replicate the analysis of this study.

